# *“At that moment, understanding the donation is crucial to make a decision”:* A qualitative study exploring the process of postmortem brain tissue donation after suicide

**DOI:** 10.1101/2021.08.10.21261758

**Authors:** Carolina Stopinski Padoan, Lucas França Garcia, Kleber Cardoso Crespo, Vanessa Kenne Longaray, Murilo Martini, Julia Carmargo Contessa, Flávio Kapczinski, Francine Hehn de Oliveira, José Roberto Goldim, Pedro Vieira da Silva Magalhães

## Abstract

**Background:** The direct examination of postmortem brain tissue can provide valuable help in refining knowledge on the pathophysiology and genetics of neuropsychiatric disorders. Obtaining postmortem consent for the donation of brain tissue after death by suicide can be difficult, as families may be overwhelmed by a violent and unexpected death. Examining the process of brain donation can inform on how the request can best be conducted, examining the existing barriers and enhancing communication, to the benefit of proxy donors.

**Methods:** This is a qualitative study, in which we employed in-depth interviews to investigate the donation process. Family members of decedents who were eligible for brain tissue donation were asked to consider the donation; irrespective of their decision, they were invited for an interview on the donation process at least 2 months after the suicide. Data collection and analyses were carried out according to a grounded theory framework, and collection, coding, and theorization occurred simultaneously.

**Findings:** Forty-one people participated in this study — 32 family members who had consented to donation and 9 who refused it. Five key themes emerged from our data analysis: the context of the families of potential donors, the invitation to talk to the research team, the experience with the request protocol, the participants’ assessment of the experience, and their participation in the study as an opportunity to heal. We arranged the main categories around 3 central stages of the experience described by participants: before the donation request, the request, and after the request.

**Conclusions:** The participants indicated that a brain donation request that is respectful and tactful can be made without adding to the family distress brought on by suicide and is viewed in a highly positive lens in the months following the event. Having an adequate understanding of tissue donation for research was crucial for satisfactory decision-making. Meeting with the donation team was considered a chance to talk to mental health professionals about suicide. Pondering brain donation was seen as an opportunity to transform the meaning of the death and invest it with a modicum of solace for being able to contribute to research.

## Introduction

The direct examination of postmortem brain tissue may greatly aid in refining the knowledge on the pathophysiology and genetics of major neuropsychiatric disorders, providing elements that are not accessible through other approaches [1, 2]. Brain tissue specimens provide researchers with the opportunity of looking into brain-specific molecules and pathways with the resolution (eg, populations of neurons) and depth of analysis that might be needed for the characterization of mental illness, especially at a time when new technologies are opening new research avenues in biochemistry and molecular biology [1,3,4]. Brain banking procedures have been continuously described and developed. For more than 3 decades, extensive efforts have been directed towards collecting well-documented specimens of postmortem brain tissue all over the world [5–7].

In psychiatry, however, brain banking is still considered to be in its infancy, with few international collections specifically devoted to research on mental illness [8–15]. Consequently, the demand for samples exceeds the supply, and researchers face critical shortages [5,14,16–18]. The further establishment of brain tissue collections depends on funding, training and use of technology, and trust by the public and potential donors [3].

Brain banking relies deeply on community attitudes, which depend on a wide range of factors. Research has so far investigated the context and cause of the donor’s death, the contextual knowledge and health literacy, the type of donation sought (whole brain or samples), the relationship with health professionals, and the method and design of the approach [11,14,19–22]. To address those peculiarities, premortem enrollment programs for brain donors have been successfully established worldwide, helping to raise awareness for the importance of human brain studies and improving donation rates through the development of a working relationship that has been satisfactory for both researchers and potential donors [3,10,23–27]. Such donations, however, are not feasible in all cases, and consent from families has to be obtained postmortem.

Suicide research is one of such special cases, in which death is unexpected and consent acquisition through a donor program is more difficult [28–33]. In these cases, the donation opportunity is usually extremely limited, and families may be overwhelmed by the recent death. The circumstances in which the request is made can also have an impact; donation requests taking place in forensic settings may be negatively influenced by the constabulary aspects and investigations [19,22,34,35]. The decision to provide postmortem consent then passes on to a next of kin, who is tasked to represent the donor’s wishes. This can be a difficult decision, considering that little information on postmortem brain donation is available to the general population and family discussions regarding this issue are probably rare, leaving the donor’s relatives unaware of his or her wishes on this matter [26,36,37]. Considering that knowing the deceased’s wishes is the strongest predictor of satisfaction with a donation, this is worrisome [38]. Also of concern is the limited timeframe for decision-making, as opposed to premortem enrollments when the donor can make his or her own decisions and have time to consider the options. In forensic pathology services, the request is usually made not long after the family receives the death by suicide notification, leaving limited time for the family to ponder the decision. Assessing the capacity of the potential donor’s next of kin to consent and giving appropriate information to secure an informed decision is of paramount importance to help families reach a harmonious decision [19,35,39,40]. Other ethical issues remain on whether families understand the process and would be satisfied to donate tissue in this circumstance. Suicide survivors may be a special population of potential donors, because of the possible shock and conflict brought on by the violent and unexpected nature of death. Notwithstanding such concerns, there is no evidence that people bereaved by suicide are offended by a request to participate in research or lack the capacity to make an informed decision [41, 42].

Brain banks of neuropsychiatric illness do not usually collect tissue after suicide [9], and the violent and unexpected nature of this death may impose the need for developing appropriate safeguard procedures for the donation process. As such, specifically studying the process of postmortem brain tissue donation to research after suicide is necessary. The major challenge for this type of research is how to raise donation rates while limiting the possibility of harm and ensuring appropriate care for potential donors and their families. A comprehensive understanding of the process of postmortem brain tissue donation is decisive not only for making this practice more effective in counteracting the decline in donation rates but also to engage the broad community in this challenge by presenting an opportunity to contribute to the health outcomes of future generations [10, 15].

For the past eight years, we have been working on the establishment of a biorepository of tissue specimens obtained from people who died by suicide [43]. As part of the donation protocol, we set up a qualitative study with in-depth interviews to investigate the donation process and its consequences. The interaction between family members and the donation team, especially the quality of communication, has a powerful and complex role in describing and predicting donation behaviors [44, 45]. A qualitative approach to these issues can clarify preferences, attitudes, and beliefs, which can be articulated through accounts that generate a comprehensive report of the interaction of those multiple influences on the donation experience [46–48], ensuring that the process is managed with sensitivity and care [49–51]. Understanding the experience with brain donation can inform on how the request process can best be conducted to the benefit of donors, while also potentially boosting donation rates. With this purpose, we present here a thorough examination of the complex process that ensues when families are presented with a request for brain tissue donation from a recently deceased close one.

## Methods

### Context of the Study

This study is part of a project for a repository of brain tissue specimens collected from people who died by suicide [43]. Family members of decedents who were eligible for brain tissue donation were asked to consider the donation. Irrespective of their decision, they received an invitation to return for an interview at least 2 months after the suicide. We designed a qualitative interview protocol to understand several issues related to the perception of the approach to tissue donation, the decision to donate, and the impact of the donation.

To gather preliminary data on opinions about brain tissue donation and on how to request family consent to donations for research, especially brain tissue, we conducted a pilot study with people who suffered from bipolar disorder and their family members because of the strong connection between severe mental illness and suicide. Participants were mostly unaware of the fact that postmortem brain tissue could be donated for research but reacted positively to the notion, appreciating the opportunity to contribute to mental health science [52].

Considering these results and based on guidelines from international biobanks, we developed a research protocol in tandem with a consent strategy in collaboration with the Bioethics Unit of the Institution [53–59]. After extensive training, including observations, role-playing, debriefing sessions, and consultation, the donation team started approaching families for consent in 2014. All approaches were made in person, in a quiet and confidential setting.

The resulting request protocol was designed to be conducted by a senior team member, usually a health professional with an MSc degree and experience with dealing with grieving families. It begins with introductions and expressions of sympathy. The donation team then presents the project and the research purposes. Autopsy procedures are described and brain harvest procedures are clarified, ensuring that the donation does not alter standard autopsy procedures, does not disfigure the donor’s body, and does not delay or impact funeral arrangements. Next, the donation of brain tissue (whole brain or sample) for research is offered to the family as an option to be considered. Families are encouraged to discuss the decision with other significant ones, and they are given space and time to reach a decision. If the donation team receives a negative response, the team thanks the family for considering the donation. If the family consents, authorization forms are signed, the answer is conveyed to the coroner’s office staff, and the brain collection process begins. After reassuring the family that their wishes will be honored, the donation team requests permission to schedule the research interview.

### Study design

We designed this qualitative study to characterize the experience of considering brain tissue donation to research after the suicide of a close one. Through their detailed narratives and prioritizing their assessment of the process, we aimed to develop a practical framework that could serve as a guide for treating families with sensitivity and respect; it would also provide families with relevant information and an opportunity to reflect on their wishes regarding brain tissue donation.

We used Strauss and Corbin’s grounded theory as framework for data collection and analysis [60], as it offers a system in which data from participants determine what is explored, how the research question should evolve to form its relevant branches, the study sample, and which literature should be explored [61, 62]. Data collection, coding, and the theorization process occurred simultaneously [63]. As such, we could be in close contact with the participants’ reactions, preferences, and ideas to improve both the design of the donation process and the protocol of the research interview. The final protocol was then constructed according to the participants’ attitudes and opinions, upholding their emotional well-being during the process.

### Recruitment and participants

Participants were adult family members of people who died by suicide and were asked to consider brain tissue donation (detailed in Longaray et al.) [43]. They were initially approached because they were present at the Medicolegal Department to attend to mandatory procedures regarding violent deaths. Because of the brain donation procedure, they also had to be approached before the necropsy was performed — all violent deaths are necropsied by law. There were no further inclusion criteria for participation in this study.

Recruitment occurred from March 2014 to November 2019. In this period, we approached 51 families while they awaited post-mortem examination procedures. We include, in this report, interviews conducted with 41 people: 32 from families that consented to donate brain tissue for research and 9 from families who refused to donate brain tissue (Fig 1). All family members were interviewed at least 2 months after the initial approach, although in some cases we were only able to interview them after several months. We conducted face-to-face or telephone interviews according to the participants’ preferences and needs. In-person interviews occurred in a specialized clinical research facility. We started the interviews with a representative of each family, but subsequent analyses informed us that the interaction between different family members played a central role in their experience, so we began inviting more than 1 family member to participate in the interviews. We conducted most interviews within a single appointment of around 150 minutes when face-to-face and of around 80 minutes when by telephone. Eighteen participants requested to be interviewed by telephone. Reasons given for not attending the in-person interview were incompatibility of schedules.

**Fig 1.**
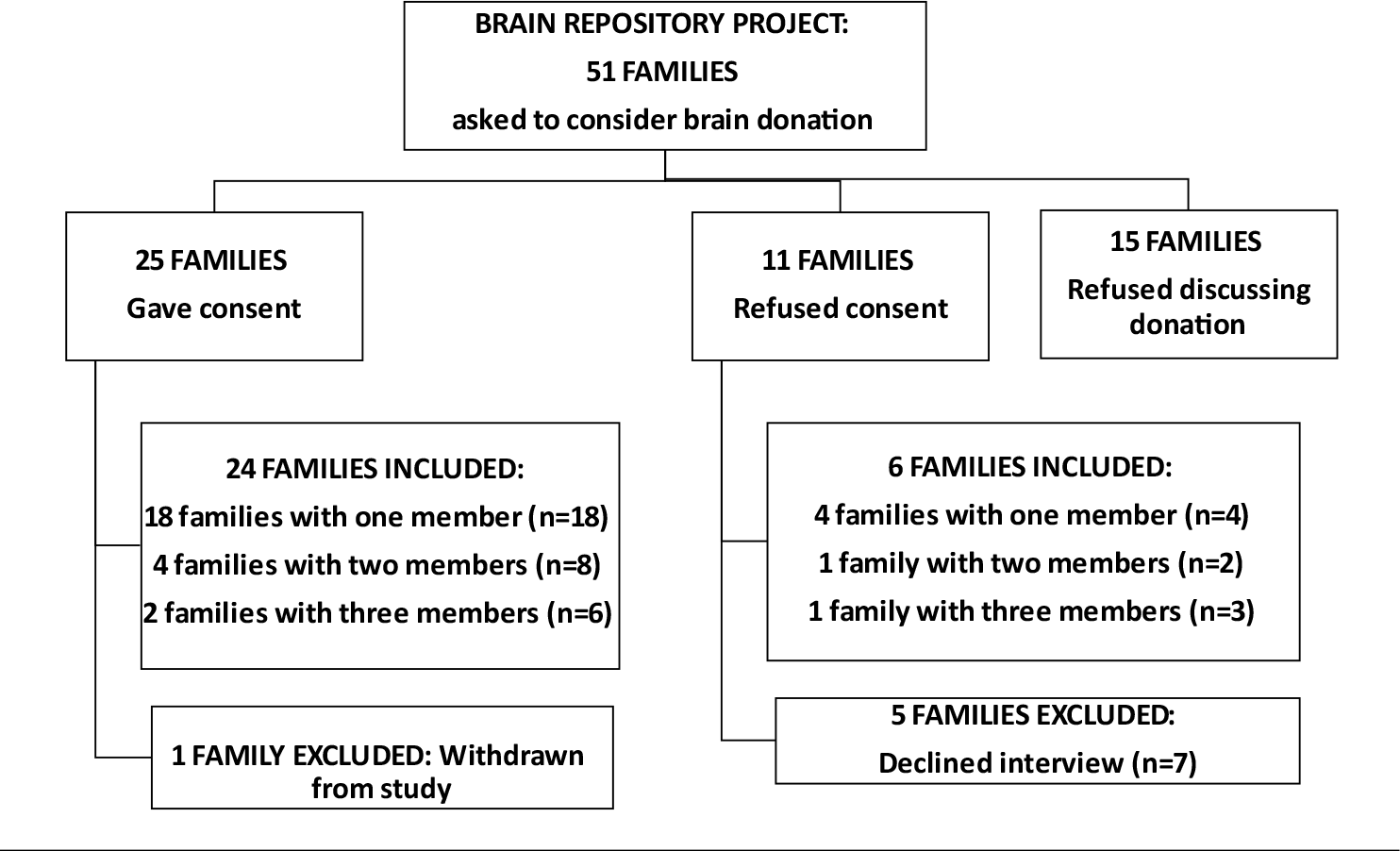
Participamt’s recruitment flow with reasons for exclusions.

We used theoretical sampling for recruitment, in two stages. In the first stage, we used a purposeful strategy to collect the maximum amount of primary data from an information-rich group with the maximum possible demographic variation, which helped us improve the interview guide and add pertinent topics [46]. Next, we sought participants’ narratives that enabled us to expand on previous understandings and further develop categories and meaning systems. In this stage, we also sought discrepancies and negative cases to prevent misleading inferences and allow transferability and generalizations [64]. Recruitment ended when the analysis reached saturation of the coding themes, which meant that each thematic category was richly detailed and complex and an innovative theory could be created, with no new elements appearing in the interviews [62,63,65].

### Instrument

We explored the family members’ experiences with the donation request and their interaction with the donation team with in-depth interviews containing open-ended questions. We used open-ended questions because we wanted to build real connections with participants while they were reacting to the interview themes and their memories of the interaction [66]. We developed an interview guide that covered the participants’ descriptions of how they received and reacted to the brain donation request. Family members were also asked to retell information about the research and donation procedures they could remember by the time of the interviews. In the final section, participants were asked to evaluate their experience with the research protocol, as well as their interaction with the donation team. We started the interviews by asking, “Before our encounter, did you know it was possible to donate organs for research?” and then followed the participants’ associations [62, 67].

The guide was modified over the data collection based on the evolution of the coding process, which informed new content that required further investigation [64]. We added a section to explore the impact of the brain donation perspective at different moments, searching for the elements involved in the transformation of initial reactions into a more open attitude. Moreover, we noticed in our first interviews that donation and research participation as opportunities to heal were a recurring theme. As a result, we added a further section for examining the meaning of the donation and research participation and its relationship with the families’ process of coping.

The final interview guide covered 8 main themes: the **conditions** and **setting** in which the families received the donation request, including prior knowledge on organ donation for research and the impact of the loss; the **initial reactions** and **impact** of the invitation to talk to the donation team; the **development** of the request **process** and the **stages** that families went through; the **decision-making** process; **attitudes and feelings** throughout the process; their **satisfaction** with the decision; **evaluations** of the request protocol and donation team and suggestions for improvement; and **views** and **meanings** that families attributed to the donation and/or research participation. In our final interviews, participants were also asked to consider preliminary results and our description of the interaction with the donation team, as we wanted to have their feedback on the trustworthiness of our conclusions.

The interviews were performed by PhD-level psychologists and psychiatrists, 2 women and 2 men (CSP, PVSM, TAC, and PDG), who were trained and supervised by a mental health and bioethics team. All interviewers had experience in the mental health field and research practices, as well as in qualitative interviews.

### Data analysis

The interviews were recorded and transcribed. Interviewers also took field notes on the impressions and feelings generated by the conversation and on the emotional state of the family members. The analysis was performed according to a grounded theory framework [60].

The analysis was conducted in pairs of researchers (CSP; LFG), and coding matrices were discussed in group meetings at all phases of the coding process. The analysis was performed with the help of NVivo software, version 12, to store interviews, select fragments of text for analysis, create codes and thematic areas, reorganize interviews following matrices after a constant comparison strategy, and create coding matrices.

The analysis process followed 3 phases: open-coding, axial-coding, and selective coding [63]. In the first stage, all transcripts were read through and coded line by line. Initial coding schemes were made of concepts and ideas, subsidiary concepts, and definitions. As the interviews progressed and the analysis continued, semantic contents were differentiated, summarized, and recorded. The constant comparative method was used to determine core categories in the axial coding phase when focused codes merged into conceptual themes. In the last phase of the coding process, we were able to organize content in the form of conceptual themes that constituted the core ideas of our work. The final coding matrices were supervised by senior experts (PVSM; JRG).

### Ethics statement

The team members who met survivors of a suicide loss were alert for signs of distress. When the need for help was detected, survivors were referred to a specialized trauma clinic. We made a referral for treatment on 17 occasions. Most of the referrals were requested by the interviewee for the benefit of another family member. In 4 circumstances, active follow-up was performed with those families to ensure medical assistance was being received. Experienced psychologists or psychiatrists from our research group made telephone calls every 3 weeks to monitor any signs of significant worsening, as well as to guarantee that medical or psychological assistance was being delivered. In this research project, there was no report of severe adverse effects caused by either donation requests or the research interview. The project was approved by the local Research Ethics Committee. In addition to providing written consent, participants also gave verbal consent at the end of each interview. We report the findings according to current Consolidated Criteria for Reporting Qualitative Research (COREQ) guidelines [68].

## Results

### Sample characteristics

Forty-one people participated in this study — 32 were from families that had consented to the donation. Participants were men and women aged 18 to 84 years; some were the key decision-makers regarding brain donation, while others assisted in the process (see Table 1).

**Table 1.** Characteristics of informants (n=41)

| Characteristics of informants | N |
| --- | --- |
| Gender |  |
| Female | 29 |
| Male | 12 |
| Age | From 18 to 75 |
| Relationship to deceased (potential donor) |  |
| Parents | 5 |
| Spouses | 9 |
| Siblings | 8 |
| Offspring | 5 |
| Second-degree relatives | 8 |
| In-laws | 5 |
| Close friend | 1 |
| Decision regarding donation |  |
| Participants from families that agreed | 32 |
| Whole Brain donation | 16 |
| Brain Tissue Sample | 16 |
| Participants from families that refused | 9 |
| Decision maker |  |
| Key decision-maker | 22 |
| Auxiliary in decision-making | 19 |
| Marital Status |  |
| Single | 10 |
| Married | 20 |
| Divorced | 4 |
| Widowed | 7 |
| Education |  |
| Elementary school | 12 |
| High school | 13 |
| College and above | 16 |

### Findings

We arranged the main categories found in data analysis around 3 central stages of the experience, as described by participants: before the donation request, the request, and after the request. Five key themes emerged: (1) the context of families of potential donors, (2) their initial reactions; (3) their experience with the request protocol, (4) appraisal of the research experience; and (5) participation in the study as an opportunity to heal. Further coding resulted in the identification of 17 related subcategories (Fig 2).

**Fig 2.**
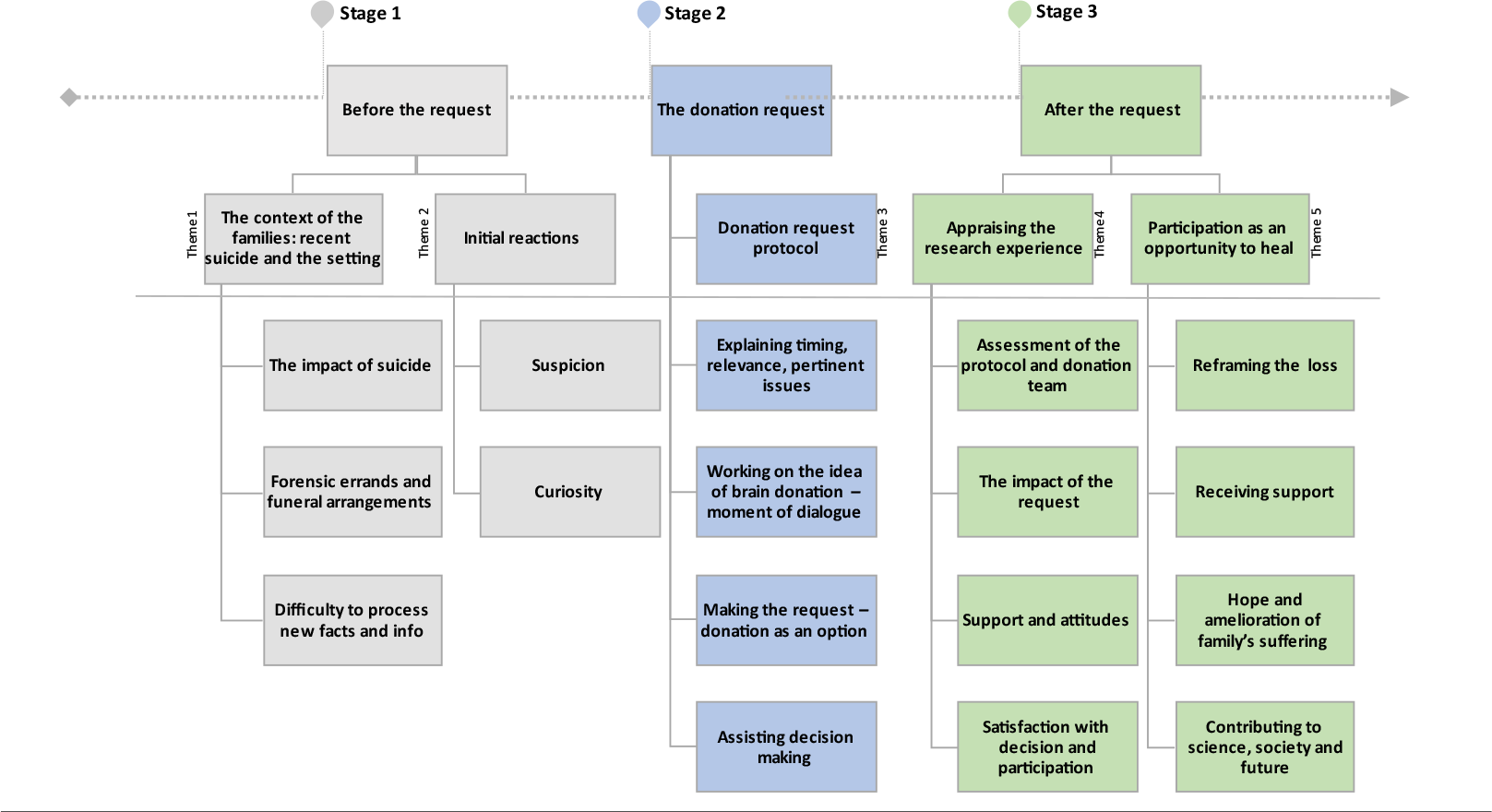
Conceptual map - the experience of brain donation for research.

### Stage 1 — Before the request

#### Theme 1: The context of families of potential donors — recent suicide and the

The first contact with the bereaved families took place while they waited on forensic authorities to release the body for funeral arrangements in a common area. At first, the violent and shocking characteristics of the suicide along with the constabulary aspects of the coroner’s office were deemed to be obstacles for understanding the study.

> *“(Suicide) is terribly hard on you. The situation and the place itself, the coroner’s office, it all makes you feel weak like you are losing your senses.” (Inf. ID 31)*

Family members emphasized suicide as an incomprehensible and disruptive phenomenon that can hinder the processing of new information.

> *“It’s not easy to think clearly because all your senses are affected by that place. You do not have good reasons to be there.” (Inf. ID 31)*

> *“We were all very disturbed at that moment, so it took us a while to fully understand the research and the donation and the safety of the procedure.” (Inf. ID 32)*

The legal actions related to the suicide, along with funeral arrangements, were painful and demanding. Nevertheless, they did not consider the donation request had made things more difficult.

> *“All that scenario, all those practicalities are awful. Autopsy. Seeing the body. Thinking about death. Talking to you about the brain was one of those things we had to do. But what was really disturbing was the suicide, not the donation.” (Inf. ID 9)*

#### Theme 2: Initial reactions

The first contact with the family was an invitation to talk privately. At this stage, suspicion and confusion emerged in some narratives. Participants associated such feelings with the events of the day and the setting.

> *“Everything that was happening to us was bizarre. So, all it came to my mind when you came to talk was bad news.” (Inf. ID 26)*

> *“They approached us with caution. We felt like it was an invitation to talk so they could tell us about this research and ask for help.” (Inf. 37)*

A few participants were concerned about being asked to disclaim personal information about the deceased.

> *“Well, I agreed to talk, but… I did not know what they wanted to talk about. I imagined all sorts of things. What if they found something in his body?” (Inf. ID 31)*

### Stage 2 —The request

#### Theme 3: The donation request protocol

Addressing the families’ needs and connecting to their pain was highlighted as a necessary step in the preparation for a conversation about brain tissue donation. Acknowledging that moment in time as a difficult one due to loss and trauma was considered of paramount importance for those bereaved to adopt an open disposition.

> *“My first impression about the request? It was not good. We were going through all that hell, and she was there wanting to talk. But then she started talking and she was overly concerned about us, about what we would think about the request. She was genuinely concerned not to disturb us any further, you know. She even said she was deeply sorry that she had to approach us at that moment in time. To me, it was fair enough.” (Inf. ID 27)*

Making it clear to families why the donation request had to happen at that moment in time was a turning point for building a relationship of mutual trust.

> *“She explained to me why they had to ask me for it (donation) at that moment in time. I got it. It was not ideal. I was very disturbed. But you do not have a suitable alternative. It is your only chance. This puts me at ease.” (Inf. 9)*

Perceiving an interest of the donation team in knowing more about suicide and mental health problems was another aspect that participants associated with building trust.

> *“I could feel they wanted answers and they were dedicated to that search.” (Inf. ID 25)*

> *“She was committed to that objective, to knowing more about suicide. It made sense to me, the utility of this research.” (Inf. ID 36)*

Participants generally lacked knowledge about organ donation for research, especially brain tissue donation. As a result, most of them were first introduced to the possibility of brain tissue donation during the approach. Nevertheless, this was not associated with discomfort or shock.

> *“I knew nothing about it. Never heard anything about it. But it was no big deal. Normal conversation considering circumstances, you know.” (Inf. ID 17)*

> *“No, it didn’t bother me to find out about brain donation. It is always better to be an informed person.” (Inf. ID 1)*

Participants recalled being encouraged to ask questions and exchange views, calling this a moment of dialog, not only about the research project but about suicide in general and mental health issues.

> *“We talked about mental health, and I enjoyed getting to know about brain studies. She answered my questions. Everything she told me then was relevant, especially for someone who was going through suicide in the family.” (Inf. ID 14)*

For most participants, talking about autopsy procedures and brain harvesting was not considered a distressing topic; they wished to understand technical aspects and deemed it positive to be able to ask the researchers for explanations.

> *“The way he explained to me how the autopsy is conducted was quite instructive. It gave me relief. We imagine terrible things.” (Inf. ID 29)*

Participants’ narratives revealed that the request for brain tissue donation was presented to them as an option to be made after they had sufficiently contemplated the issue.

> *“In the end, the researcher offered us the possibility of donating her brain to the study. It was a chance to participate and contribute. She made it clear that we could say yes or no. It was not something that I felt I was obliged to do.” (Inf. 17)*

Participants believed that having the space and time to ponder the options helped them reach their decision. They also deemed it positive to be encouraged to exchange views with significant others, even with relatives that were not present if that was the case. The last point they highlighted was the importance of having assistance with difficult questions. For most, there was fear of damaging the donor’s body, that retrieval could delay autopsy procedures, and about the purpose of the donation.

> *“At first, it was hard for me to consider the donation. But then we talked. She explained to me that they would use it for studies, for research. When research is the point at stake, you must think about it. And I remember thinking that I would like to help with this research. I would like to contribute to this mission.” (Inf. ID 29)*

> *“The way they handled our fear of having his body damaged made us feel at ease. And they gave us time to decide and did not push it. They said we could decide whatever was best for the family. It was an option that was offered to us.” (Inf. 15)*

The final report on feelings and reactions towards the donation approach at this phase of the interaction was unanimously *“tranquil,” “at ease,”* and *“fine”*.

### Stage 3 — After the request: appraising the research experience and participation as an opportunity to heal

#### Theme 4: Appraising the research experience.

Families mentioned a respectful attitude, candor, credibility, clarity of speech, politeness, and professionalism as the most important positive qualities of the donation team.

> *“They were very professional, very correct. You could see they were serious.” (Inf. ID 30)*

> *“The donation proposal was well conducted in my opinion. Everything was explained to me, the study and what they were trying to investigate. You could see they had it organized.” (Inf. ID 25)*

Participants who were invited to comment on the structure of the interaction in our final interviews felt that this organized the experience by addressing emotions first, including a moment for those in need to share their stories, facilitating comprehension. This was also considered a demonstration of empathy and care towards families and of respect for the deceased.

> *“That was an extremely complicated situation that we were facing, losing someone we love to suicide. It was turmoil for everybody. But he delivered the donation prospect very calmly, step by step. First, he told us all about the research, and then he discussed the donation and why it was so important. In the end, we were at ease.” (Inf. ID 27)*

Participants considered that understanding the information was a vital aspect of the experience that gave them the confidence to decide. The use of written material with adequate terminology was believed to aid the process, and it helped those who consented to explain the procedures to the rest of the family.

> *“It (written material) helped me to explain it all to my family over the phone so they could help me decide” (Inf. 33)*

> *“At that moment, understanding the donation is crucial for someone to make a decision. If you can’t understand it, you can’t decide, because you get paralyzed by fears and misconceptions. That’s what the conversation with your people helped us with.” (Inf. 25)*

Participants reported that although they were surprised by the request, they were not further stressed or upset by it, even in the face of sudden bereavement. Being asked to consider a donation at that difficult moment in time seemed not to disrupt the families’ emotional state. Even though participants did not claim to be negatively surprised by the request when having no previous knowledge about the topic, they all stated that it made the decision harder on them. Being impacted by a sudden and violent loss together with having to ponder on something new to them was challenging.

> *“The problem of not knowing about it previously is that you are already facing a situation that you’ll never understand, the suicide. And at the same time, you do not have any knowledge about this kind of donation. It just makes it more of a challenge to process. It may put people against it (donation) just for the trouble of being unable to think clearly.” (Inf. 31)*

The negative aspects of the approach were mainly related to the forensic setting. The coroners’ office was associated with violent scenarios, violent deaths, and intrusive procedures. Families complained about unpleasant odors they assumed came from dead bodies. Waiting room areas were teeming with persons going through loss or accident, which participants recalled with distress.

> *“That place is horrible. It is nobody’s fault. But it is an aggressive setting, full of people going through some psychological trauma. And the smell… It is awfully strong and bizarre. It adds to the burden of the loss.” (Inf. 11)*

We made a final assessment of the protocol in our final interviews, sharing our conceptual model of the interaction with family members. The narratives showed that the current protocol adheres to the families’ needs.

> *“I relate to those reports. You do not have to change anything. Keep on following up with families. This is genuinely nice.” (Inf. ID 25)*

> *“I feel satisfied. I appreciate the way they approached me and handled the situation. It was a normal conversation. Clear and focused, not aggressive. No rambling on futile stuff. When you are in the middle of a disaster, you do not have time to lose. They told me their business and they asked for my opinion. Fair enough.” (Inf. ID 17)*

Family members reported feeling satisfied with the care they received from the researchers. They were also satisfied with the decision on the donation. One family member revealed being sorry for having their opinion overruled by the rest of the family, who did not agree with the donation. Another participant, the key decision-maker, mentioned that they wished they had accepted.

> *“Afterwards, I kind of regretted having said no. I think I could have questioned myself more. It’s one thing you are not expecting, having to make this decision…” (Inf. 26)*

> *“I feel sorry that the others refused it. It was quite shocking for me the way they totally rejected the idea. I tried to talk to them, make them see things more clearly, but they got upset with me. So, I gave it up. But in my opinion, they should have said yes to the donation.” (Inf. 13)*

Participants showed support for brain donation, even in the face of loss and trauma, regardless of their decision. In their opinion, the need to advance suicide prevention through mental health science justified the approach. What made them consider the request safe for people in distress was the guarantee of the right to decline. Once mourning is respected and acknowledged, they believe the donation request should be made.

> *“Keep on doing it and keep on trying to get donations. This is a way of helping mankind fight mental disease.” (Inf. 16)*

> *“This donation is a very good thing for our society because it may help to discover why people do it and you guys are trying to make us all help a little.” (Inf. ID 29)*

#### Theme 5: Participation in the study as an opportunity to heal

The interaction with the donation team had a positive effect on those participating. Collaboration with this project, whether through brain tissue donation or interviews, was an opportunity to give a different meaning to the life and death of their deceased relative. The donation was associated with a feeling that the death of their loved ones was *“not in vain,” “leaving something positive behind.”* The act of contributing to research represented a chance to mitigate the helplessness left by suicide by contributing to advance knowledge on this issue and bringing hope for a scientific breakthrough.

> *“It (the approach) kind of changed my state of mind. It took the focus off of the cruelty of that place. It gave me something positive to consider while I was there (coroner’s office) instead of just the pain. It marked my experience; it was a turning point.” (Inf. ID 33)*.

> *“It changed (the donor’s) legacy a bit. In the end, (the donor) made something good. (The donor) went all their life in this wrong path, lots of misdeeds, unhappiness. And now this good action is their last action.” (Inf. ID 1)*

Participating in research that aims to further our knowledge on suicide was considered a helpful way to deal with the urge to understand that particular suicide by transforming a personal quest into a broader action of helping future generations.

> *“It is of some relief to know that we are helping to fight this tragedy.” (Inf. ID 28)*

> *“This research gave me hope. Because I realized that what I was facing, losing a loved one like that, is a widespread problem that causes extensive damage to lots of families. And something must be done to prevent it. And the fact that I could help, I will have this for the rest of my life.” (Inf. ID 33)*

Participants considered the conversation with the donation team as a chance for a much-needed dialogue about suicide and mental health with qualified professionals that would not have happened otherwise so close to their loss. Being able to express thoughts and feelings on their terms and not having to conceal the very worst of what they were going through was beneficial for most families.

> *“It was an opportunity for us to receive support from someone that had skills to help us deal with what we were facing. Her ability to address our suffering helped us a lot.*

> *Because if you know nothing about suicide, you may as well harm those who are in pain.” (Inf. ID 17)*

> *“Nobody wants to talk about this stuff, death, suicide, mental health. All they want is to gossip about the one who died. That’s why I appreciated having the chance to talk to you guys.” (Inf. ID 25)*

## Discussion

People participating in this qualitative study indicated that a brain tissue donation request can be made without adding to the very significant family distress brought on by suicide. A request that is respectful and tactful, acknowledging their suffering, can overcome initial suspicions and is viewed in a highly positive lens in the months following a suicide loss. Having an adequate understanding of donation for research emerged as the crucial process for satisfactory decision-making. Meeting the donation team was additionally considered a chance to talk to a mental health professional about suicide. Pondering brain donation was also seen as an opportunity to transform the meaning of the death and invest it with a modicum of solace for being able to contribute to research.

There has been some debate on whether recently bereaved families can be expected to make truly informed decisions in face of the recent loss to suicide [35,38–40,69,70]. Suddenly bereaved individuals are reportedly at higher risk for mental health issues, which are frequently associated with difficulty understanding and processing new information [42,71–73]. As a result, the capacity of grieving individuals to freely give consent has been questioned and must be assessed [35]. The endeavor of brain banking in psychiatry has to face the challenge of dealing with potentially vulnerable individuals. Nonetheless, the stigma that surrounds suicide and mental health issues, which may perpetuate a sense of secrecy for fear of discrimination, has led the research community towards the careful inclusion rather than outright exclusion of vulnerable individuals [70,73–75]. The key feature here is that vulnerable groups have a right to participate in research, and vulnerability signals the need to develop appropriate safeguard actions to empower and promote their agency in a research context [5,74,76]. While we did show here both difficulty processing information and a degree of initial suspicion, participants indicated that these could be mitigated by a donation team that made an effort to acknowledge and respect the pain they were in [77, 78]. Confirming previous research data, this study suggests that organizing the request in progressive stages can aid the person in understanding the research and donation information, retain that information in mind, and use it to exchange views with family members and the donation team as part of the decision-making process without undue pressure [35,51,7]. We believe that decisive aspects to increase a person’s ability to process donation information occur in the early stages of the approach. Initial reactions of shock and suspicion, which are frequent and expected after an organ donation request, represent an opportunity for donation teams to demonstrate positive regard and empathy, which are predictors of a positive encounter even at a difficult time [38,79– 81]. As seen in previous work, we found that conceding that the moment of the request is problematic and acknowledging the inconvenience can help suddenly bereaved individuals to separate the distress regarding the death from the discussion on brain donation [22,38,82–84].

The immediate experiences of suicide-bereaved individuals can be exceptionally traumatic [8]. They may have found the deceased’s body; they may have dealt with police officers at their home investigating the death scene; and finally, families have to not only try to understand what happened, but also try to explain it to extended family and friends. At the same time, the ones at the morgue have to keep other relatives updated on the legal situation [35,86–88]. Some participants of our study revealed how these experiences triggered feelings of anxiety regarding the autopsy results or what the research might reveal about the donor. We believe the fundamental aspect here is to keep a vigilant eye for such a possibility, reiterating the donation team’s commitment to the best interests of the participants.

Families were generally unknowledgeable of the possibility of brain tissue donation for research, which is something we explored before [52]. Brain donation is a sensitive topic among the general public, for whom relevant and culturally sensitive information is scarce [89]. Indeed, having little contextual knowledge and health literacy can influence donation, not only by affecting rates of consent but also by making it a more difficult personal decision due to lack of familiarity with the theme [16,24,36,90]. Understanding how brain donation works and why it is fundamental for mental health research was considered a vital step towards carefully considering a donation. When presented with the option, they felt that having time to discuss with the family and feeling that was an opportunity they could decline were useful in reaching a measured and harmonious decision. Even if being surprised by the brain donation request was not associated with negative reactions in this study, all participants claimed they would prefer to know more about it before the request because the knowledge gap was considered an obstacle to processing information and reaching a decision that best represented their wishes. Reiterating the recommendations from brain donation literature, the implementation of active outreach awareness campaigns and culturally relevant educational protocols is needed globally, not only to address the current decline in post-mortem donations but to ensure that families have a positive experience with the donation request [17,23,38,50,91,92].

Providing clear and accessible explanations about the necessity and benefits of this specific field of research for mental health science is a necessary step towards effective communication with potential donors [2,86,93]. Details on the autopsy, brain harvest, impact on the donor’s body, and funeral arrangements are better processed if delivered repeatedly [24]. Moreover, in this study, we observed that when the donation team clarified the reasons why they were committed to the research, people could relate to their work, and this helped form a relationship of mutual trust. Previous work on donation requests demonstrates that trust enables people to express themselves fully and ask questions [39], establishing a two-way conversation that engages family members in the process, which is vital for pondering donation options.

Negative feelings and uncertainties can be changed by information given by donation teams at this stage of the approach [94]. Additionally, the initial inclination to instinctively recoil from the matter and decline may be mitigated by allowing for an examination of their underlying motives and drives [93]. This process often results in a decision where the facts prevail [93, 95]. Most importantly, when donation teams act to aid families in deciding what best represents their wishes, the approach is more frequently associated with positive feelings and non-maleficence [22,92,96,97]. Working on the idea of brain donation relies on frequently checking in to confirm the understandings, but most importantly, the misunderstandings of the family members regarding what the donation entails [16]. It is vital to allow people to interject and raise issues, as well as to encourage discussion among family members. By doing so, donation teams open space for a needed discussion regarding fears, misconceptions, and expectations, while offering clarifications that can aid in making the decision [19, 98]. What we identified here, with the aid of the participants’ narratives, is that donation teams must educate people before they can ask them to consider brain donation for research, and this implies working beyond the delivery of information. Possessing the tools to comprehend what is being asked makes families engage in conversation, which gives them time to explore the request and make their decision in association with their values and feelings.

The participants were widely satisfied with their decision to take part in the study. Not only did they demonstrate an understanding of the need for obtaining brain tissue for research and felt it did not add to their emotional burden, but they also mentioned the participation as an opportunity to see the deceased more positively. For some, the opportunity to talk to a mental health professional about suicide was also helpful. When participants were asked to review their experience with the research, most of them referred to the thoughtfulness of the approach and the importance of the research with donated brains. This finding supports previous research, showing that the manner of the request influences families’ perceptions of the donation experience, how families see the approach as a whole, and the level of information they receive [14,22,24,38,80,92,99,100].

There is a growing body of literature showing that donation for research has the potential to be rewarding for bereaved families [16,22,38,50,79,80,83,100–102]. Making an impact on other people’s lives and having the potential to help others in the future are frequently considered sources of emotional and practical comfort that bereaved families derive from donating to research [38, 50]. Since most families that are faced with the possibility of brain donation for research consider it a good thing and a source of comfort and hope, they state that the opportunity to donate to research should be granted to all families as a right [22, 102]. Accordingly, the participating families were unanimously supportive of the enterprise of brain donation regardless of their decision to donate. As feedback, they asked the donation team to continue offering the possibility of brain donation and research participation to all bereaved families in the future. Fortunately, and in line with more than a few previous studies, suddenly bereaved families are not further distressed by being asked to consider brain donation for research [22,36,38,51,80,83,93,100,102]. Negative reactions towards the donation are reportedly tied to requests that are not culturally tailored when the conversation about donation does not meet families’ needs, and the care for donor relatives is not a priority [26,50,77,103].

Particularly for those consenting to the donation, the experience had the power to reframe negative aspects of the suicide and the deceased, adding meaning to the experience. This finding is frequently reported in other studies, in which the act of donating is related to a better capacity to cope and make sense of the death, facilitating the bereavement process [21,22,104]. When specifically considering the feelings of hopelessness and guilt, which are very much common in survivors of a suicide loss, a donation can assume great significance, providing a sense of agency for doing something worthwhile, as a last act they could do for the deceased and for all those who suffer from mental health problems [50,105,106].

Postmortem donation requests can also offer an initial point of contact between health professionals and suddenly bereaved individuals. Previous research indicated that initial grief reactions such as shock and denial can make it hard for family members to ask or recognize the need for help and counseling [80]. Particularly worrying is that support is crucial for survivors of suicide loss, yet they are less likely to receive it immediately and are more likely to report a delay in receiving it than people bereaved by sudden natural causes of death [106, 107]. Talking about donation with a healthcare professional can increase access to health care and raise awareness about the risk of complicated grief symptoms [108]. Health professionals strongly influence people’s experiences with the donation process; this contact can be a valuable source of hope and support, one that can even facilitate a referral to specialized health care when needed.

Brain tissue donation for research can awaken a sense of personal duty, as described by Lin and colleagues. People depict the donation as a way to help others and society in general, which served to facilitate their grieving process and offered a degree of comfort. But the opportunity to benefit from the donation conversation needs not be limited to donating families. Two key aspects were identified by the participants of this research as accounting for a benevolent and positive encounter with the donation possibility, irrespective of the final decision. The first one was having the donation team showing appreciation for their attention to the conversation and recognizing their efforts to deal with a scientific novelty at a difficult moment in time. It made people feel respected and valued. The other aspect involved offering the donation as an option to be made after enough pondering and consultation within the family and only when, in addition to representing their wishes, it was not considered to make things more difficult for the rest of the family. Feeling comfortable to decline made people state that brain donation for research should be offered to all families as a right. Receiving personal attention and support from a health care professional, even in cases where the donation was not authorized, was described as beneficial to the grieving process, and a source of hope and relief.

The impact of suicide can be a barrier to processing new information, but we suggest here it can also serve as motivation for learning more about research that intends to further our knowledge on mental health. The initial suspicion can come along with curiosity, and this disposition can and should be addressed, imposing the need for a confident requester. Quality communication is a two-way path, which means that this initial disposition, despite being emotionally charged, is a point of entry for a much-needed clarification of factual information. This is why donation conversations cannot be rushed, and families have to feel comfortable expressing real concerns and expectations about what a brain donation entails.

Looking back at the experience with the donation prospect, following the family’s lead is what matters the most (Fig 3). Some people will need to talk about their emotions and what happened with the deceased, while others will prefer to focus on the research and donation aspects. Adjusting the approach accordingly, without detracting from all the important information needed to secure an informed decision, is indispensable. This flexibility can only be achieved when the manner of the request is well planned and organized. Using the feedback from those who were previously approached elevates not only the manner of the request but the donation team’s confidence in the protocol. In the Box (Fig 4), we highlight several key points we believe can be drawn from this investigation.

**Figure 3.**
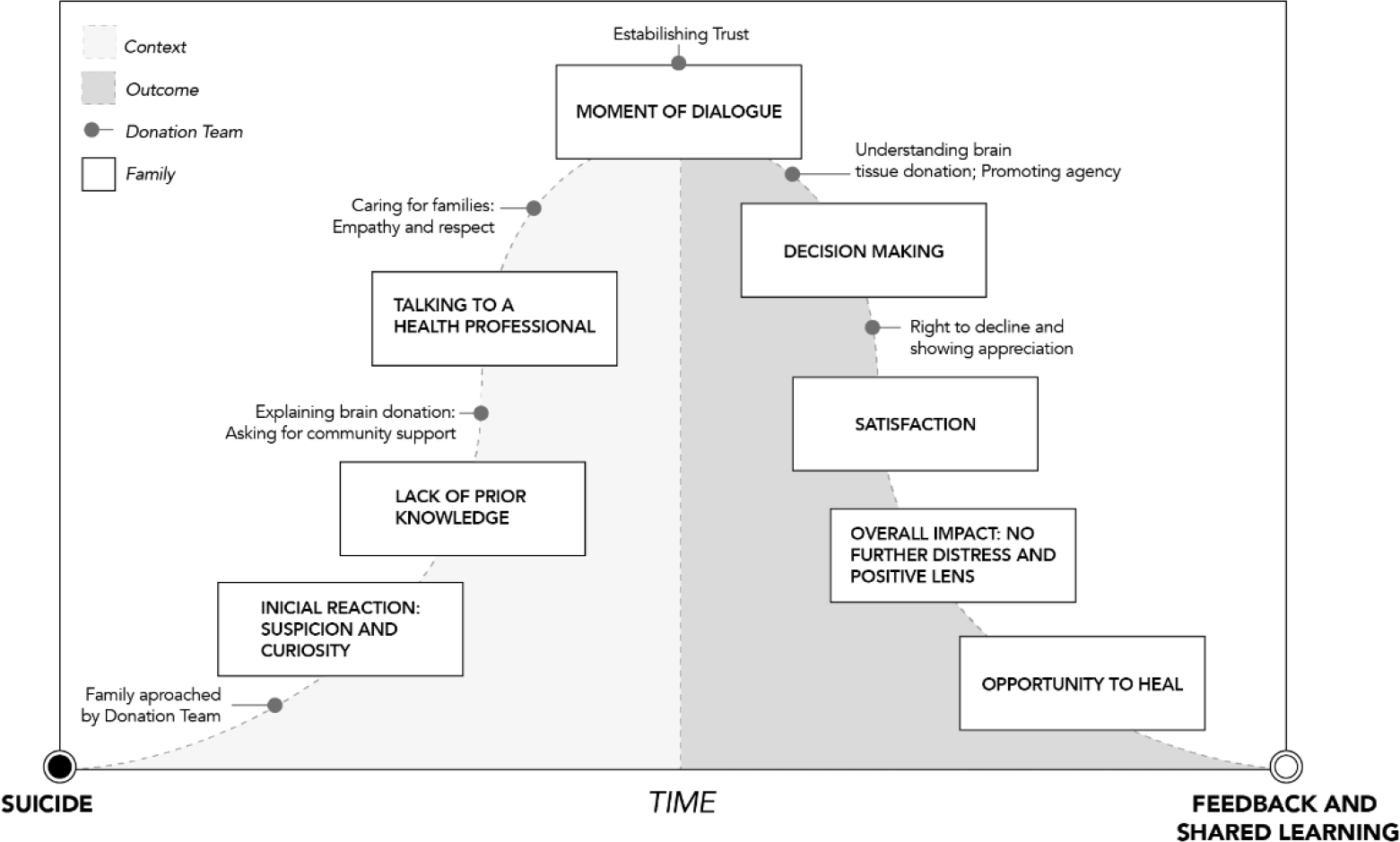
Grounded theory on stages leading to brain tissue donation for research after suicide.

#### Key elements for a thoughtful request for brain donation for research

- Those asking must be **well trained** and **confident**
- Give **personal attention** and make a real **connection** to family’s suffering.
- Remember to be **open** and **honest** about what you are asking for and the reasons why it is so important.
- Before you ask, educate, and **work on the idea of brain donation**. Emphasize facts and Procedures.
- Provide an informational sheet to take home, including contact details in case any further question arises.
- Follow the **family’s pace** and adjust your protocol accordingly.
- Check understandings AND **misunderstandings** regarding donation.
- Provide accurate information to **aid in making the decision**.
- Encourage **discussion within family members**. Give them **time** to ponder.
- Offer donation as an **option** to be made after sufficient pondering time.
- Adjust your process for the request with the **help of the feedback** from those that were previously approached.
- Be mindful that an adequate request can be anxiety-provoking, but it also has the **potential to be extremely rewarding** when it best represents the family’s values and wishes, creating an opportunity to heal.

Even if the reactions to the approach described here were overwhelmingly positive, some families would not consent to be interviewed. As such, we were unable to ascertain their possible reactions and whether they would be troubled by the approach. It was our impression that the most troubled families were precisely those that refused any contact, and their outlook on donation and research participation in general may be completely different from what we found here. We chose not to fully explore the details of the decision-making process here and expect to report them soon elsewhere. Another limitation refers to the study sampling procedure. Although the qualitative design was fundamental for an in-depth examination of families’ experiences, our sample was correspondingly small; this made our findings not representative of all recently bereaved families. Qualitative studies can, nevertheless, point to themes that could be more broadly investigated; future surveys with larger samples should be helpful to verify the findings of this study. Naturally, these findings may not apply to brain donation requests conducted premortem, and this issue has been approached elsewhere [27, 109].

## Conclusion

While we present here a wealth of novel findings on people’s experiences with brain donation and interactions with the donation team, we also advance a theory grounded on these experiences. We believe such theories are vital to developing an approach to tissue donation for research that builds on trust, to help grieving families make the best-informed decision based on their values. Our study suggests that brain donation in the context of a recent suicide can be made in an informed and respectful manner, and families are overwhelmingly satisfied with their decisions. Our findings also point to the perceived benefits of postmortem donation, which could be even more meaningful for those bereaved by suicide.

## Data Availability

The data that support the findings of this study are available from the corresponding author, upon reasonable request.

## Acknowledgments

We thank all the families who participated in this work; the managing, teaching, and research sections, and the morgue staff at the Medicolegal Department; Instituto Geral de Perícias do Rio Grande do Sul; and the Bioethics and Science Ethics Research Laboratory, Hospital de Clínicas de Porto Alegre (HCPA). We acknowledge the invaluable assistance in data collection by Dr. Taiane de Azevedo Cardoso, Dr. Pedro D. Goi and Dr. Angelita Maria Ferreira Machado Rios.

This study was funded by Conselho Nacional de Desenvolvimento Cientıfico e Tecnologico (CNPQ), Brazil financed in part by the Coordenação de Aperfeiçoamento de Pessoal de Nível Superior - Brasil (CAPES) - Finance Code 001. Professor Magalhaes is supported by a National Council for Scientific and Technological Development – CNPq productivity fellowship.

## Consolidated criteria for reporting qualitative studies (COREQ): 32-item checklist

Developed from:

Tong A, Sainsbury P, Craig J. Consolidated criteria for reporting qualitative research (COREQ): a 32-item checklist for interviews and focus groups. *International Journal for Quality in Health Care*. 2007. Volume 19, Number 6: pp. 349 – 357

| No. Item | Guide questions/description | Reported on Page # |
| --- | --- | --- |
| **Domain 1: Research team and reflexivity** |  |  |
| *Personal Characteristics* |  |  |
| 1. Interviewer/facilitator | Which author/s conducted the interview or focus group? | 13 |
| 2. Credentials | What were the researcher's credentials? E.g. PhD, MD | 13 |
| 3. Occupation | What was their occupation at the time of the study? | 13 |
| 4. Gender | Was the researcher male or female? | 13 |
| 5. Experience and training | What experience or training did the researcher have? | 8-9, 13 |
| *Relationship with participants* |  |  |
| 6. Relationship established | Was a relationship established prior to study commencement? | 8, 9, 10, 13, 14, 19, 20 |
| 7. Participant knowledge of the interviewer | What did the participants know about the researcher? e.g. personal goals, reasons for doing the research | 12, 13, 14, 15, 26 |
| 8. Interviewer characteristics | What characteristics were reported about the interviewer/facilitator? e.g. Bias, assumptions, reasons and interests in the research topic | 12- 13, 22-25 |
| **Domain 2: study design** |  |  |
| *Theoretical framework* |  |  |
| 9. Methodological orientation and Theory | What methodological orientation was stated to underpin the study? e.g. **grounded theory**, discourse analysis, ethnography, phenomenology, content analysis | 8-14 |
| *Participant selection* |  |  |
| 10. Sampling | How were participants selected? e.g. purposive, **convenience**, consecutive, snowball | 10-12, Fig1. |
| 11. Method of approach | How were participants approached? e.g. face-to-face, telephone, mail, email | 10-12 |
| 12. Sample size | How many participants were in the study? | 10, 11, 15 Table1 |
| 13. Non-participation | How many people refused to participate or dropped out? Reasons? | Fig1 |
| *Setting* |  |  |
| 14. Setting of data collection | Where was the data collected? e.g. home, clinic, workplace | 8 - 10 |
| 15. Presence of non-participants | Was anyone else present besides the participants and researchers? | 12 |
| 16. Description of sample | What are the important characteristics of the sample? e.g. demographic data, date | 11, 12, Table 1 |
| *Data collection* |  |  |
| 17. Interview guide | Were questions, prompts, guides provided by the authors? Was it pilot tested? | 12-13 |
| 18. Repeat interviews | Were repeat interviews carried out? If yes, how many? | 12-13 |
| 19. Audio/visual recording | Did the research use audio or visual recording to collect the data? | 13 |
| 20. Field notes | Were field notes made during and/or after the inter view or focus group? | 13 |
| 21. Duration | What was the duration of the interviews or focus group? | 11 |
| 22. Data saturation | Was data saturation discussed? | 11-12 |
| 23. Transcripts returned | Were transcripts returned to participants for comment and/or correction? | 10, 13, 22-25 |
| **Domain 3: analysis and findings** |  |  |
| *Data analysis* |  |  |
| 24. Number of data coders | How many data coders coded the data? | 13-14 |
| 25. Description of the coding tree | Did authors provide a description of the coding tree? | 13-14, 16-17, Fig 2 |
| 26. Derivation of themes | Were themes identified in advance or derived from the data? | 13-14, 16-17, Fig 2 |
| 27. Software | What software, if applicable, was used to manage the data? | 13-14 |
| 28. Participant checking | Did participants provide feedback on the findings? | 10, 13, 22-25 |
| *Reporting* |  |  |
| 29. Quotations presented | Were participant quotations presented to illustrate the themes/findings? Was each quotation identified? e.g. participant number | 17-27 |
| 30. Data and findings consistent | Was there consistency between the data presented and the findings? | 22-25, 27-37, Fig 3 |
| 31. Clarity of major themes | Were major themes clearly presented in the findings? | 16-17, Fig 2, Fig 3, Box |
| 32. Clarity of minor themes | Is there a description of diverse cases or discussion of minor themes? | 27-37 |

**S1_Fig.**
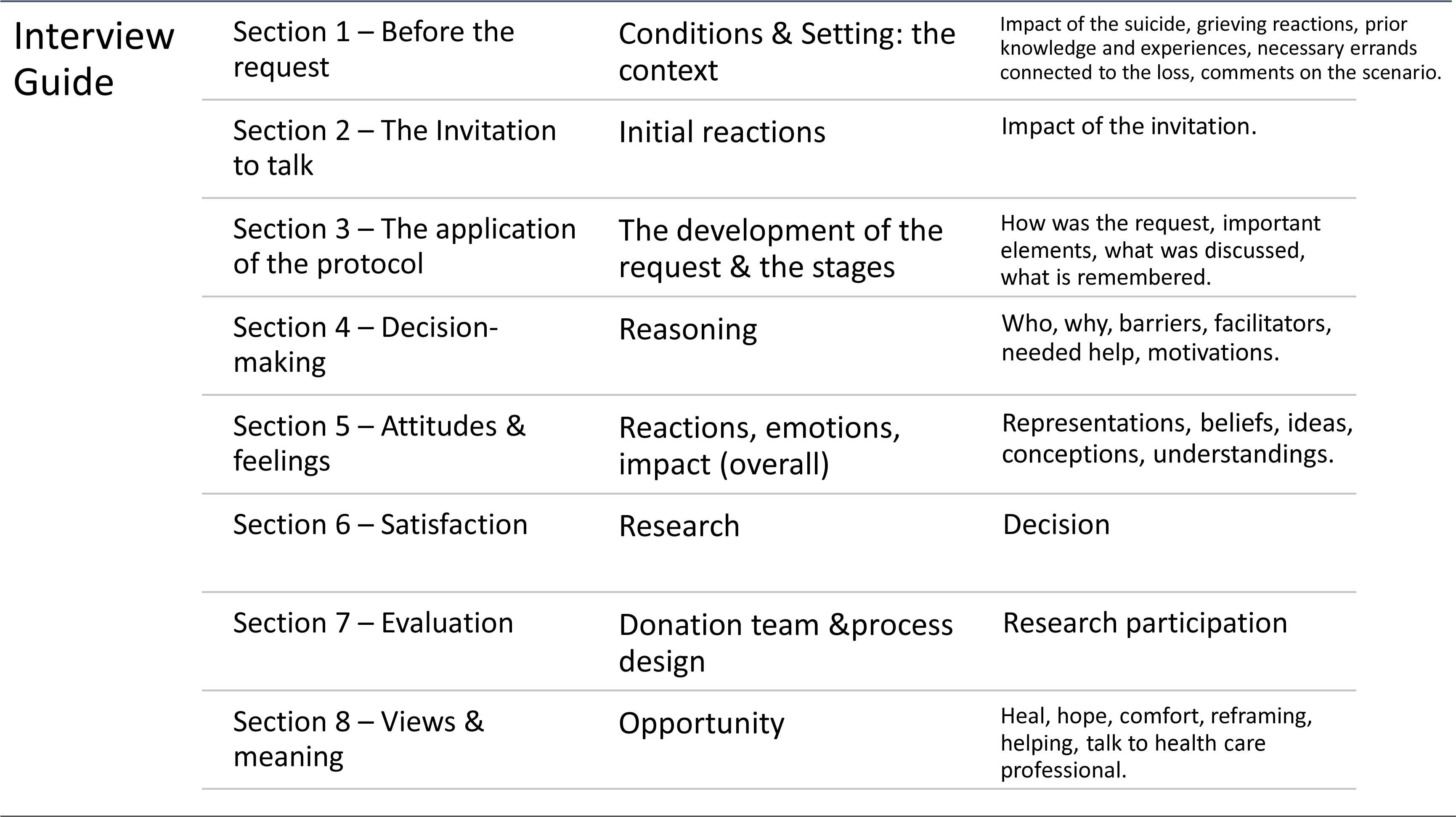
Interview guide.

